# Surgical modification of deep brain stimulation lead trajectories substantially reduces RF heating during MRI at 3 T: From phantom experiments to clinical applications

**DOI:** 10.1101/2022.12.22.22283839

**Authors:** Jasmine Vu, Bhumi Bhusal, Joshua Rosenow, Julie Pilitsis, Laleh Golestanirad

**Affiliations:** Department of Biomedical Engineering, McCormick School of Engineering, Northwestern University, Evanston, IL, USA; Department of Radiology, Feinberg School of Medicine, Northwestern University, Chicago, IL, USA; Department of Neurosurgery, Feinberg School of Medicine, Northwestern University, Chicago, IL, USA; Department of Neurosciences & Experimental Therapeutics, Albany Medical College, Albany, New York, USA

**Keywords:** deep brain stimulation (DBS), magnetic resonance imaging (MRI), radiofrequency heating

## Abstract

**Introduction:** Radiofrequency (RF) induced tissue heating around deep brain stimulation (DBS) leads is a well-known safety risk during magnetic resonance imaging (MRI), resulting in strict imaging guidelines and limited allowable protocols. The implanted lead’s trajectory and its orientation with respect to the MRI electric fields contribute to variations in the magnitude of RF heating across patients. Currently, there are no consistent requirements for surgically implanting the extracranial portion of the DBS lead. This produces substantial variations in clinical DBS lead trajectories and hinders RF heating predictions. Recent studies showed that incorporating concentric loops in the extracranial trajectory of the lead can reduce RF heating, but the optimal positioning of the loop remains unknown. In this study, we systematically evaluated the RF heating of 244 unique lead trajectories to elucidate the characteristics of the trajectory that minimize RF heating during MRI at 3 T. We also presented the first surgical implementation of these modified trajectories and compared their RF heating to the RF heating of unmodified trajectories.

**Methods:** We performed phantom experiments to assess the maximum temperature increase, ΔT_max_, of 244 unique lead trajectories. We systematically interrogated the effect of three characteristics related to the extracranial portion of the lead trajectory, namely, the number of concentric loops, the size of the loops, and the position of the loops on the skull. Experiments were performed in an anthropomorphic phantom implanted with a commercial DBS system, and RF exposure was generated by applying a high-SAR sequence (T1-weighted turbo spin echo dark fluid pulse sequence, B_1_^+^_rms_ = 2.7 μT). Test-retest experiments were conducted to assess the reliability of measurements. Additionally, we determined the effect of imaging landmark and perturbations to the DBS device configuration on the efficacy of low-heating lead trajectories. Finally, recommended modified trajectories were implanted in patients by two neurosurgeons and their RF heating was characterized in comparison with non-modified trajectories.

**Results:** Our search protocol elicited lead trajectories with ΔT_max_ from 0.09 – 7.34 °C. Interestingly, increasing the number of loops and positioning them near the surgical burr hole—especially for the contralateral lead—substantially reduced RF heating. Trajectory specifications based on the results from the phantom experiments were easily adopted during the surgical procedure and generated nearly a 4-fold reduction in RF heating.

**Discussion/Conclusion:** Surgically modifying the extracranial portion of the DBS lead trajectory can substantially mitigate RF heating during MRI at 3 T. Simple adjustments to the lead’s configuration can be readily adopted during DBS lead implantation by implementing small concentric loops near the surgical burr hole.

## Introduction

Since the introduction of deep brain stimulation (DBS), it has provided remarkable therapeutic benefits to patients with movement disorders and other neurological diseases [1]–[7]. During DBS, electrical stimulation is delivered to specific subcortical targets in the brain via implanted electrodes that are connected to a programmable implantable pulse generator (IPG), which is placed in the clavicle region and connected to the leads via subcutaneous extensions.

Magnetic resonance imaging (MRI) is a highly versatile neuroimaging modality with potential benefits for patients with DBS devices. It is well-suited for postoperative monitoring, target verification and localization of the electrodes [8]–[10], as well as monitoring the functional effects of stimulation on affected brain networks [11]–[16]. The need for MRI is steadily growing, and an estimated 66-75% of patients with DBS implants will require an MRI exam within 10 years following device implantation [17].

Radiofrequency (RF) induced heating is the primary limiting factor for performing MRI exams on patients with DBS devices. Localized tissue heating occurs since the implanted DBS lead behaves like an antenna when coupled with the electric field of the MRI transmit coil [18]–[20]. This phenomenon, known as the *antenna effect*, increases the specific absorption rate (SAR) of RF energy deposited in the tissue surrounding the lead’s tip. Due to safety concerns associated with RF heating, DBS manufacturers have established stringent device-specific guidelines for MRI protocols. To date, most neuroimaging procedures are limited to those at a field strength of 1.5 T in a horizontal closed bore scanner. Heating-related thresholds remain conservative, with B1+rms < 1.1 μT or whole-head SAR < 0.1 W/kg—30 times below the FDA’s limit for scanning in the absence of implants [21]. These guidelines limit routine clinical MRI for patients with DBS devices.

Efforts dedicated towards mitigating MRI-induced RF heating for patients with DBS systems have increased in recent years. These contributions include modifying the material and design of DBS leads [22], [23], introducing novel MRI head coil technology to induce a region of low electric field that coincides with the implanted lead’s trajectory on a patient-specific basis [24]–[30], and potential application of ultra-high-field [31] and vertical open-bore scanners which have different orientations of the magnetic and electric fields [32]–[34]. Although promising, the clinical utility of these approaches is still limited as they require changes to existing DBS or MRI technology or methodologies.

Surgical modification of the extracranial portion of the DBS lead trajectory is an alternative method for reducing RF heating by minimizing coupling of the leads with the MRI electric fields without altering the DBS system or MRI methodology [35], [36]. Currently, precise surgical guidelines are established for implanting the intracranial trajectory of the lead. That is, the entry point on the skull and the angle of insertion are predetermined to target the intended brain structure. On the contrary, there is a lack of similar guidelines for implanting the extracranial portion of the lead as this does not contribute to the therapeutic effects of DBS for the patient. This results in substantial variations in the extracranial lead trajectories across patients, which in turn leads to highly variable—and unpredictable—RF tissue heating [37]. Baker et al. first introduced the concept of manipulating the DBS lead trajectory into a looped configuration using a trajectory formation device [38]. More recently, one proposed modified trajectory configuration involves surgically shaping the extracranial portion of the DBS lead into concentric loops near the respective burr-hole [35], [37]. The effect of the concentric loops was predominant for the contralateral DBS leads, demonstrating an 18-fold reduction in the maximum 1g-averaged SAR in simulations with a loaded transmit body coil [35]. However, the optimal positioning of the loops remains unknown. Other studies that have implemented loops near the surgical burr-hole fixated the lead trajectory throughout their studies [39]–[41]. Pertinent RF heating-related parameters including loop dimensions and the trajectory’s position in the phantom were not fully evaluated.

The goal of this study was to perform the first large-scale, systematic study to identify characteristics of extracranial DBS lead trajectories that minimize RF heating during MRI at 3 T. We evaluated the RF heating of 244 unique lead trajectories in an anthropomorphic phantom with a commercial DBS system. Lead trajectory parameters of interest included the trajectory topology (i.e., the number of concentric loops), location of the trajectory relative to the skull, and the size of the loops. We also evaluated the RF heating of the lead trajectories under varying IPG configurations and imaging conditions to determine the robustness of our experimental methods and the low-heating trajectories. Finally, we demonstrated the feasibility of translating low-heating lead trajectories from phantom experiments to DBS patients at our institutions.

## Materials and Methods

### Lead trajectory parameter optimization

We systematically examined the effect of three parameters of extracranial lead trajectory on RF heating: the diameter of the concentric loops (2.5-4.5 cm with 0.5 cm increments), position of the loops on the skull that are surgically and anatomically feasible, and the number of concentric loops (1-3 loops) (shown in Fig. 1. B). These parameters were selected as prior studies demonstrated their nontrivial effects on the variations in the magnitude of RF heating across trajectories [35], [38], [39], [42], [43]. To establish precise replication of the intended trajectories at the MRI scanner and to ensure that experiments were reproducible, we created 3D models of the lead trajectories and the anthropomorphic phantom in a CAD tool (Rhino 7.0, Robert McNeel & Associates, Seattle, WA) which allowed for exact measurements. We then translated the intended trajectories to a commercial DBS system using 3D printed trajectory guides (shown in Fig. 1). Evaluated trajectories are shown in Figures 2 and 3 and are categorized by trajectory parameter.

**Figure 1:**
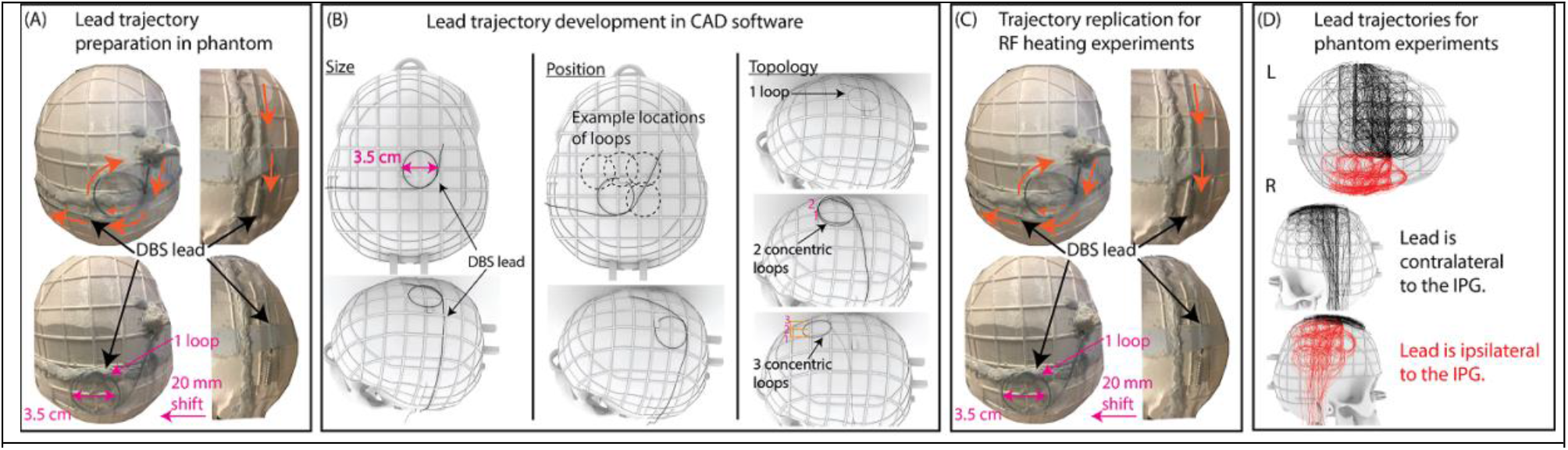
Extracranial DBS lead trajectory development process. (A) Lead trajectories were prepared on the 3D-printed skull phantom to determine feasibility the of the trajectories. (B) 3D models of all trajectories were created in a CAD tool. Trajectory parameters included the diameter of the loops, position of the loops on the skull, and topology. (C) Trajectories were replicated with a commercial DBS system. Experimental trajectories matched their digital counterparts. (D) Superposition of all the trajectories evaluated in this study. There were 150 trajectories for the lead contralateral to the IPG and 94 trajectories for the lead ipsilateral to the IPG.

**Figure 2:**
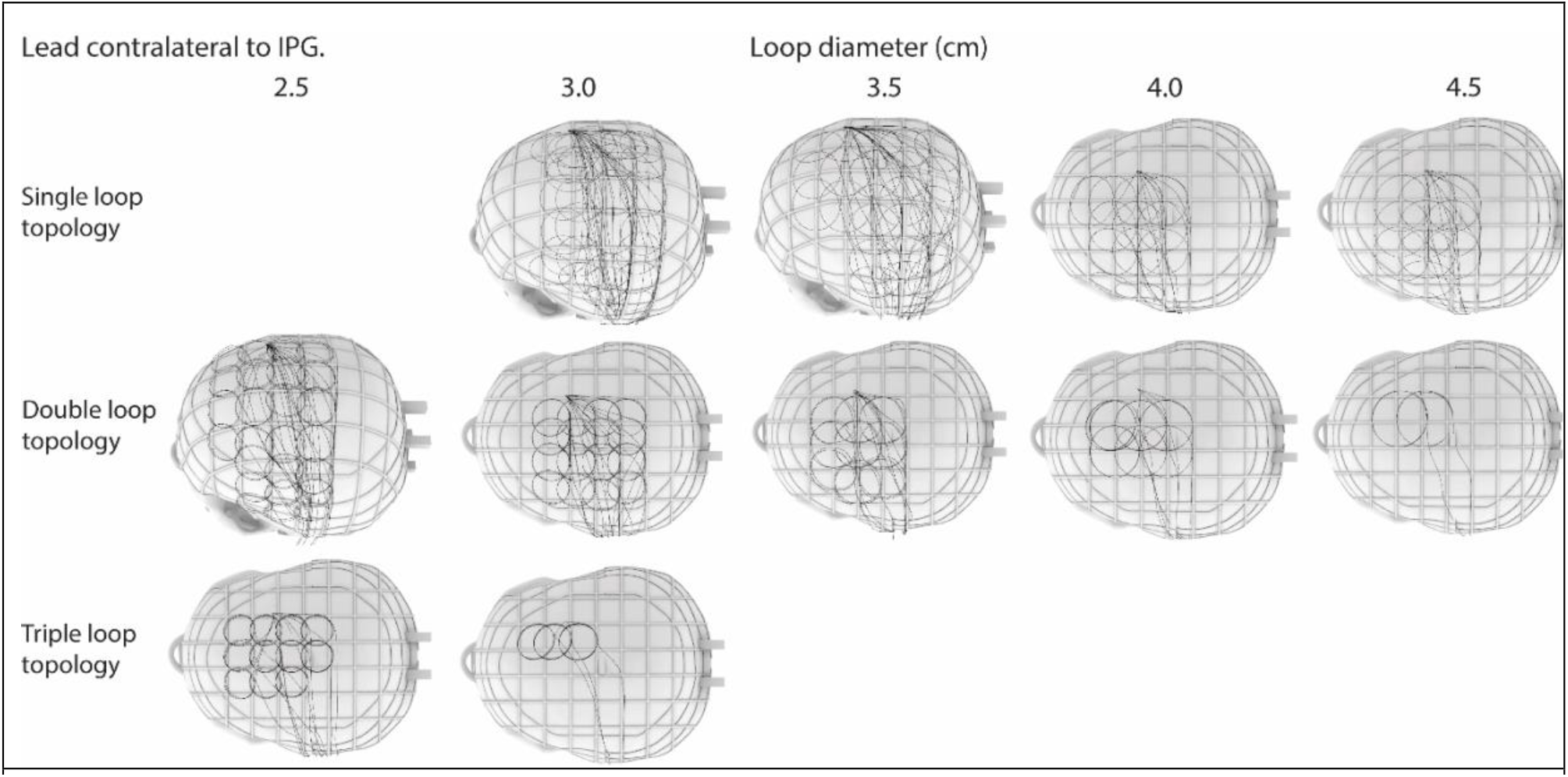
Evaluated lead trajectories categorized by loop topology and loop dimension. There were 150 trajectories for the lead contralateral to the IPG.

**Figure 3:**
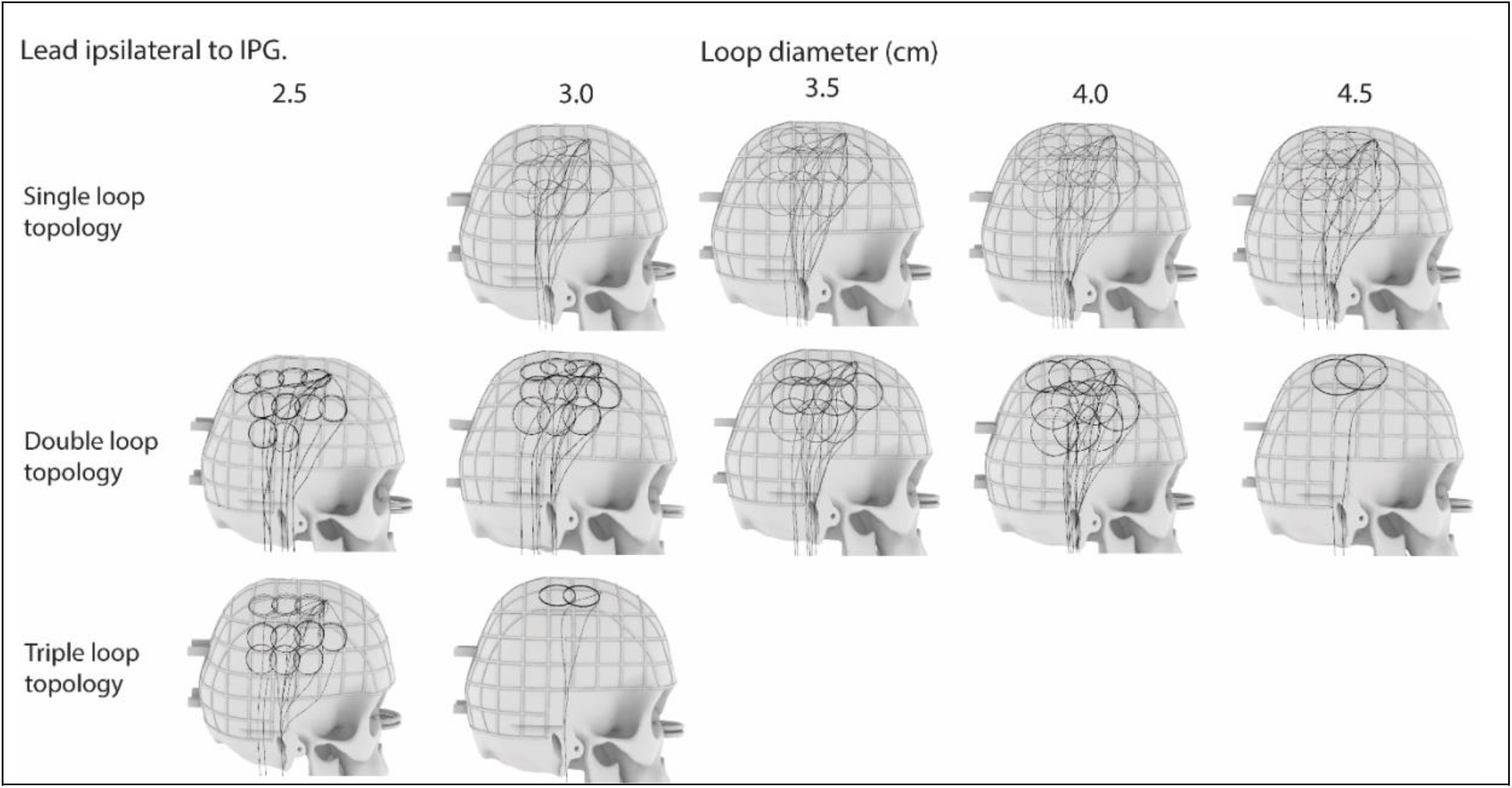
Evaluated lead trajectories categorized by loop topology and loop dimension. There were 94 trajectories for the lead ipsilateral to the IPG.

### Phantom fabrication

While the majority of RF heating studies of active implants are performed with box-shaped phantoms, the electric field distributions—and consequently SAR—are substantially different in a box-shaped phantom compared to human body models [44]. Thus, we fabricated an anthropomorphic phantom based on CT images of a patient with a DBS device (shown in Fig. 4. A) [45]. Details of the phantom design and fabrication are provided elsewhere [45]. To ensure that the skull was anatomically representative, we compared the size of the skull against the head dimensions—namely the maximum head length, maximum head breadth, and distance from the most superior point on the head to an axial plane intersecting the eyebrows—and geometries of 53 other patients with DBS devices (shown in Fig. 4. B). The maximum head breadth, length, and distance of the skull was 141 mm, 185 mm, and 91 mm, respectively (additional details are included in the Supplementary Materials). Similarly, the mean ± standard of the maximum breadth, length, and distance was 157.3 ± 8.7 mm, 198.9 ± 11.6 mm, and 100.0 ± 11.4 mm, respectively, across the 53 patients. The skull also included a grid with 20 mm spacing to assist with specific positioning of the DBS lead trajectory (shown in Fig. 4. A).

**Figure 4:**
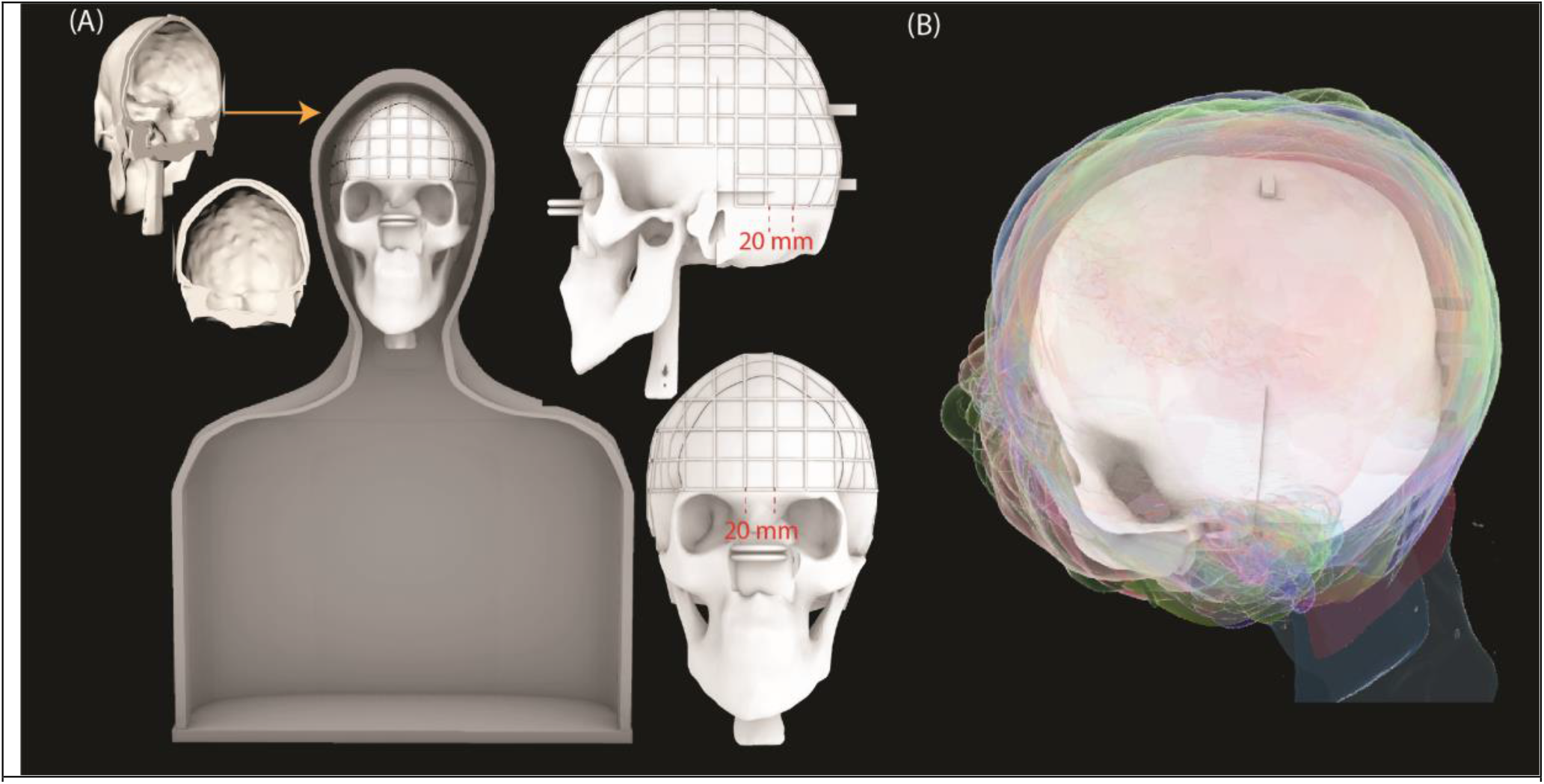
(A) 3D models of the phantom shell and the skull. A grid with 20 mm spacing was designed directly on the skull to assist with DBS lead trajectory positioning. (B) The size and geometry of the skull was compared against the head sizes of 53 other patients with DBS systems (patient heads are displayed as transparent outlines).

### Experimental setup

A total of 244 measurements were performed to determine DBS lead trajectories that minimized RF-induced heating. Experiments were performed with an Abbott full DBS system (40 cm lead model 6172, 50 cm extension model 6371, and Infinity-5 IPG). The lead was implanted in the right hemisphere (i.e., entry point on the right side of the skull) with the location and angle of insertion similar to what is done for targeting the subthalamic nucleus (STN) (shown in Fig. 5). The IPG was placed either on the right pectoral region to represent ipsilateral lead trajectories (N = 94) or in the left pectoral region representing contralateral lead trajectories (N = 150). The skull was filled with an agar-based gel (σ = 0.47 S/m, *ε*_r_ = 78) prepared by mixing 32 g/L of edible agar (Landor Trading Company, gel strength = 900 g/cm^2^), 5 g/L of sodium benzoate (Sigma Aldrich), and saline solution (1.55 gNaCl/L). The rest of the phantom was filled with 18 L of saline solution (σ = 0.50 S/m, *ε*_r_ = 80) mimicking the conductivity of average human tissue. The temperature increase, ΔT_max_, was measured in the gel surrounding the lead-tip using fiber-optic temperature probes (Osensa, Burnaby, BC, Canada resolution = 0.01 °C). An additional temperature probe was placed in the body of the phantom away from the DBS device to ensure that the temperature did not increase elsewhere in the phantom. Experiments were performed in a 3 T Siemens Prisma MRI scanner using the body transmit coil (Siemens Healthineers, Erlangen, Germany) (shown in Fig. 5). The phantom was placed in the head-first, supine position and oriented such that the eyebrows/tip of the DBS lead was at the scanner’s isocenter. RF exposure was generated with a T1-weighted turbo spin echo dark fluid pulse sequence (TR = 2750 ms, TE = 8.2 ms, FA = 170°, acquisition time = 381 seconds, B_1_^+^rms = 2.7 μT).

**Figure 5:**
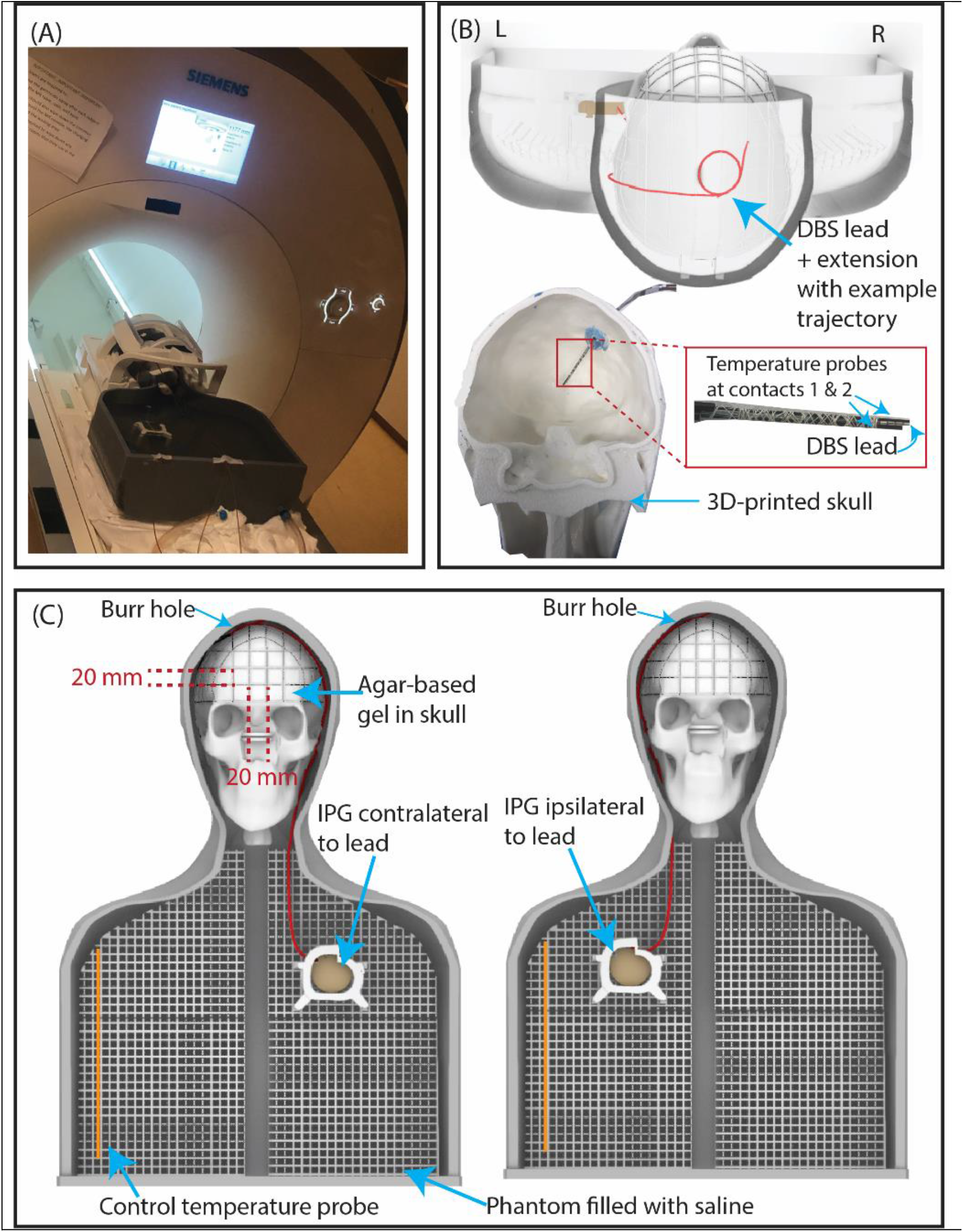
(A) Experimental setup at a 3 T Siemens Prisma scanner. (B) 3D rendering of an example lead trajectory in the 3D-printed phantom. Temperature probes were attached to the DBS lead. (C) Phantom setup for configurations where the lead was contralateral and ipsilateral with respect to the IPG.

### Test-retest analysis

To evaluate the reliability of our measurements, we repeated experiments with 18 randomly selected trajectories from which nine were contralateral and nine were ipsilateral with respect to the IPG. These trajectories represented those that generated RF heating in the top and bottom 20% as well as those in between these thresholds. We calculated the Intraclass Correlation Coefficient (ICC) and the 95% confidence intervals in the R software in RStudio (version 4.2.1) (https://www.r-project.org/) based on a two-way mixed-effects model, single rating, and absolute-agreement [46].

### Additional analysis on the effect of patient positioning, imaging landmark, and IPG configuration

Experiments were repeated for the 18 randomly selected trajectories incorporated in the test-retest analysis to determine the effect of slight changes in patient positioning on RF heating. Originally, the eyebrows/tip of the DBS lead was located at the scanner’s isocenter. The second and third patient positions were ± 20 mm superior and inferior to the eyebrow level (shown in Fig. 6.) as different indications for brain MRI may require different imaging locations.

**Figure 6:**
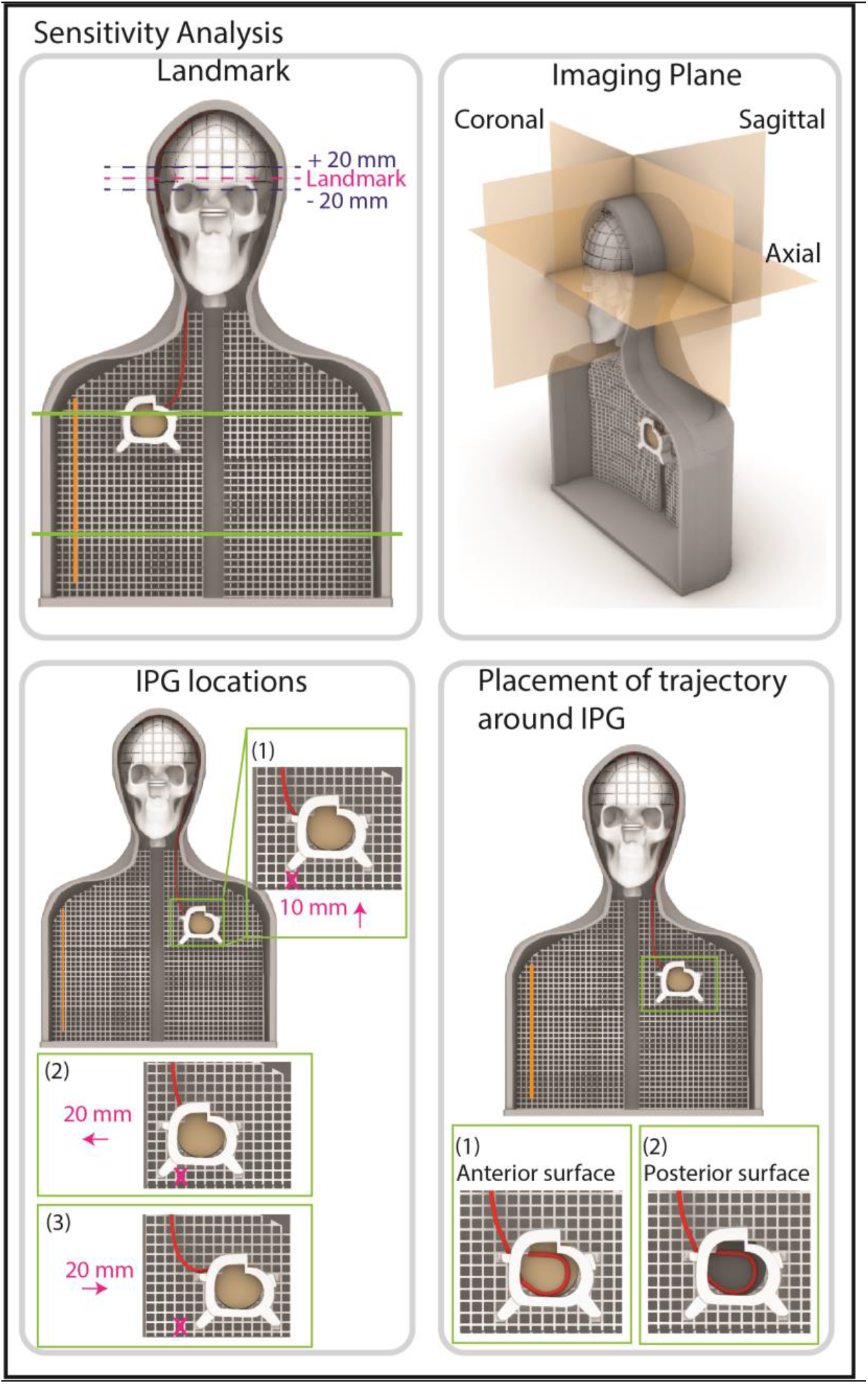
The effect of imaging and implant-related parameters including the landmark (brain, chest, and abdomen), imaging plane and its phase encoding direction, and IPG location and configuration on ΔT_max_ was evaluated. Axial slices were originally acquired. The placement of the trajectory on the anterior surface of the IPG was the original configuration.

Additionally, we performed experiments with another group of 12 randomly selected trajectories (six contralateral and six ipsilateral with respect to the IPG) that represented the top and bottom 20% of generated RF heating for imaging in the chest and abdominal regions (shown in Fig. 6). These experiments were performed to demonstrate that the trajectories which exhibited minimum heating for brain imaging did not induce excessive RF heating when the patient was positioned to image other parts of the body (i.e., chest and abdomen).

Finally, experiments were repeated for six randomly selected trajectories (three contralateral and three ipsilateral with respect to the IPG) to assess the effect on RF heating when perturbing the imaging plane, location of the IPG, and the trajectory of the extension around the IPG [47]. Experiments were performed for acquisition of coronal (right-left and feet-head phase encoding) and sagittal (anterior-posterior and head-feet phase encoding) slices (shown in Fig. 6). The location of the IPG was translated 10 mm superior to its original position and 20 mm in the medial and lateral directions (shown in Fig. 6). The trajectory of the excess extension around the IPG was originally placed anterior to the surface of the IPG; however, the trajectory was placed posterior to the surface of the IPG in the additional experiments (shown in Fig. 6). These experiments were performed to assess the robustness of trajectory modification in reducing RF heating when subjected to various perturbations that may occur in practice.

We calculated the absolute difference in ΔT_max_ between the original configuration and each configuration where a change was made to the landmark, imaging plane, or an IPG-related configuration. This difference in ΔT_max_ is indicated as |ΔT_max2_-ΔT_max1_|.

### Surgical implementation of modified lead trajectories in patients

To assess if low-heating trajectories could be readily adopted during DBS surgery, two neurosurgeons (J. R. at Northwestern Memorial Hospital and J. P. at Albany Medical Center) were instructed to implement low-heating trajectories in their patients undergoing DBS surgery. For some patients (J.P), we used curved mayo scissors passed posterior and to the left of the incision which were opened to their widest position to create a pathway for a coiled lead to be inserted. We then coiled the lead upon itself in 2-3 concentric circles at the burr-hole before passing the rest of the lead toward the temporal lobe where it would be later connected to the extension (shown in Fig. 7. A). For other patients (J.R.), the scalp was routinely elevated circumferentially from the skull to facilitate a tensionless closure using a periosteal elevator. The lead was then anchored with the burr hole ring, clip, and cap. If this lead was to be connected to an ipsilateral IPG, the electrode tip was secured in its protective cap and then passed under the scalp to the location of a previously determined extension incision that would be used during the IPG implant stage. If this lead was to be connected to a contralateral IPG, the lead was passed to an extension with a blocking pin that had been placed at the time of the IPG implantation and the two were connected. In either case, the electrode was then coiled into 3 loops which were placed under the scalp posterior to the main linear incision (shown in Fig. 7. B).

**Figure 7:**
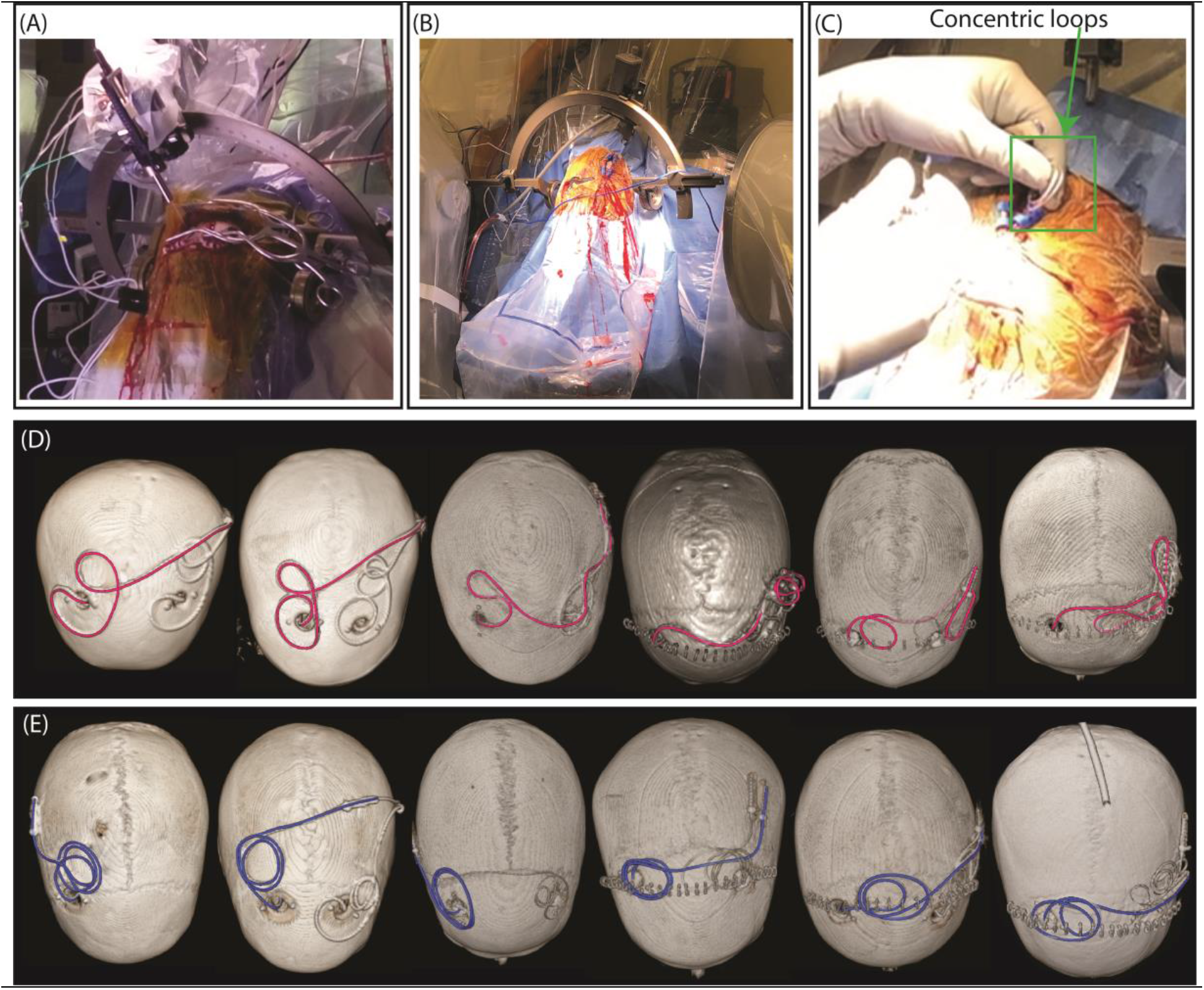
(A) DBS implantation surgery at Albany Medical Center. (B) DBS implantation surgery at Northwestern Memorial Hospital. (C) Concentric loops implemented near the surgical burr hole similar to the phantom experiments. 3D surface rendered views of CT images of patients with (D) unmodified (highlighted in magenta) and (E) modified (highlighted in blue) DBS lead trajectories that were replicated during phantom experiments.

Following the procedures, lead trajectories were segmented from postoperative computed tomography (CT) images using 3D slicer (http://slicer.org) and processed in the Rhino CAD tool. Models of the trajectories were 3D-printed to help with trajectory replication of a commercial DBS device during phantom experiments. Furthermore, experiments were performed with lead trajectories from the same neurosurgeons prior to receiving instructions on implementing low-heating trajectories to compare the effectiveness of surgical lead modification. Retrospective use of patients’ imaging data for the purpose of modeling was approved by Northwestern Memorial Hospital and Albany Medical Center’s institutional review boards.

## Results

### RF heating measurements

Leads that were contralateral with respect to the IPG had higher heating compared to leads that were ipsilateral to the IPG based on a two-sample one-tailed t-test (p = 2.44 × 10^−23^). The mean ± standard deviation of ΔT_max_ was 3.44 ± 1.93 °C with a range of 0.24-7.34 °C for leads contralateral with respect to the IPG (shown in Fig. 8. A). For leads that were ipsilateral to the IPG, the mean ± standard deviation of ΔT_max_ was and 1.26 ± 1.17 °C with a range of 0.09-4.75 °C (shown in Fig. 8. B). All loops were coiled in the clockwise direction.

**Figure 8:**
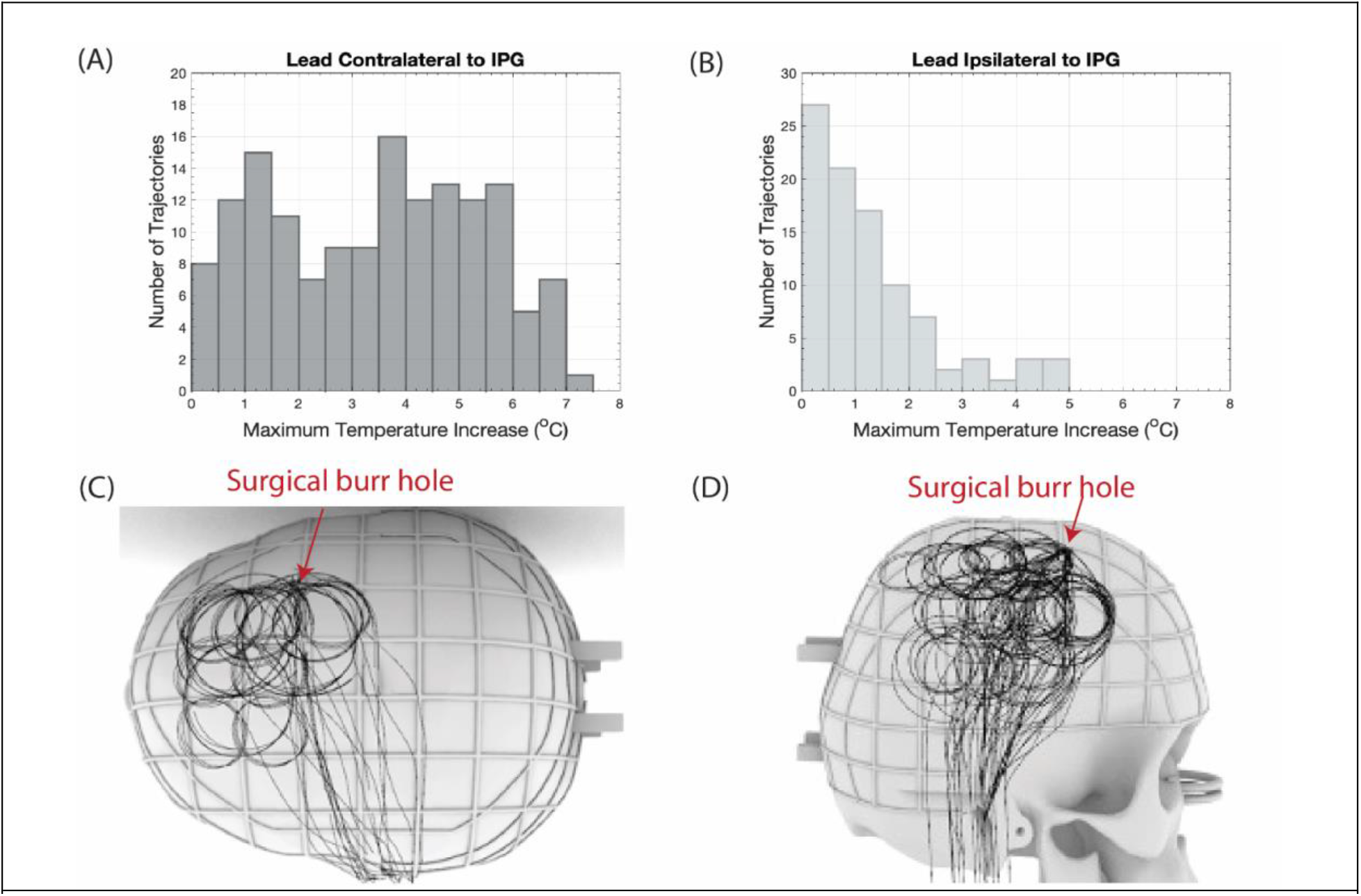
Distribution of ΔT_max_ for all lead trajectories (A) contralateral and (B) ipsilateral with respect to the IPG. Superposition of low heating trajectories where ΔT_max_ < 1°C to illustrate the anatomical position of these trajectories (C and D).

### Effect of spatial orientation

Low-heating trajectories had concentric loops positioned closer to the surgical burr hole (shown in Fig. 8. C and D). This trend was especially apparent for leads that were contralateral to the IPG. For lead trajectories where ΔT_max_ < 1 °C, 16 out of 20 and 27 out of 48 trajectories were within 40 mm radially of the surgical burr hole for contralateral and ipsilateral leads, respectively. The impact of the anatomical location of the concentric loops was consistent regardless of the number of concentric loops and the size of the loops.

### Effect of number of loops

The mean ± standard deviation of ΔT_max_ was 4.99 ± 1.03 °C, 2.07 ± 1.14 °C, and 0.73 ± 0.40 °C for single, double, and triple loop configurations, respectively, for leads contralateral to the IPG (shown in Fig. 9. A-C). Similarly, the mean ± standard deviation of ΔT_max_ was 1.79 ± 1.39 °C, 0.79 ± 0.82 °C, and 1.19 ± 0.56 °C for single, double, and triple loop configurations, respectively, for leads ipsilateral to the IPG (shown in Fig. 9. D-F). As the number of concentric loops increased from one to three in contralateral trajectories, ΔT_max_ decreased (correlation coefficient, r = -0.83).

**Figure 9:**
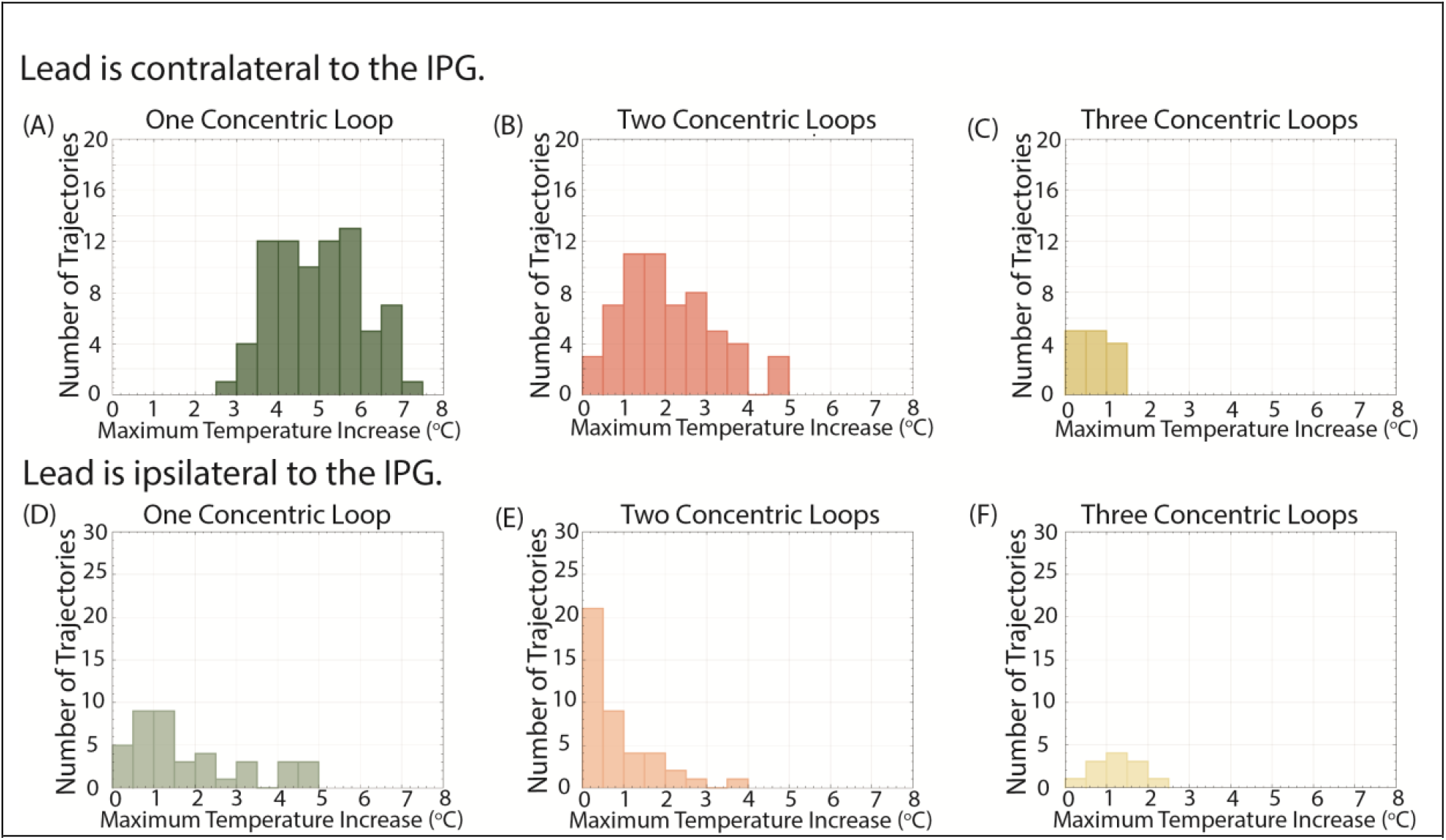
Distribution of ΔT_max_ categorized by the number of concentric loops (A-C and D-F).

### Effect of loop size

The diameter of the loops in the contralateral trajectories did not correlate with ΔT_max_ (r = 0.17). Table 1 provides the mean ± standard deviation of ΔT_max_ for each loop size for leads contralateral (shown in Fig. 10. A-E) and ipsilateral (shown in Fig. 10. F-J) with respect to the IPG.

**Table 1:** Effect of loop size on ΔT_max_

| Diameter of loop(s) (cm) | Mean ± standard deviation of $\Delta T_{\max}$ ($^{\circ}\text{C}$) | |
| --- | --- | --- |
|  | Leads contralateral to IPG | Leads ipsilateral to IPG |
| 2.5 | $2.11 \pm 1.22$ | $0.71 \pm 0.59$ |
| 3.0 | $4.04 \pm 1.98$ | $1.52 \pm 1.62$ |
| 3.5 | $4.11 \pm 2.01$ | $1.43 \pm 1.22$ |
| 4.0 | $3.35 \pm 1.95$ | $1.46 \pm 1.09$ |
| 4.5 | $3.38 \pm 1.36$ | $1.11 \pm 0.65$ |

**Figure 10:**
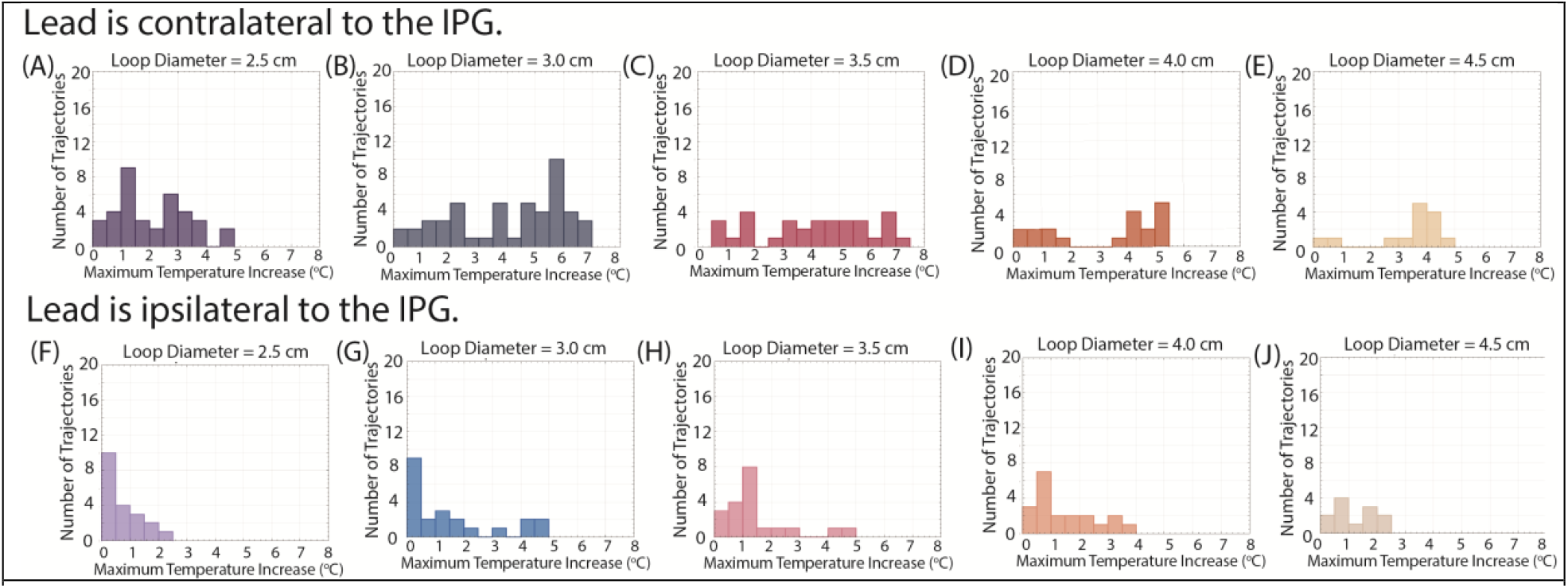
Distribution of ΔT_max_ categorized by the diameter of the concentric loops (A-E and F-J).

### Test-retest measurements

The repeated experiments demonstrated excellent reliability where the ICC = 0.96 with a 95% confidence interval of 0.91-0.99. Figure 11. A. shows the ΔT_max_ values for the 18 test-retest experiments.

**Figure 11:**
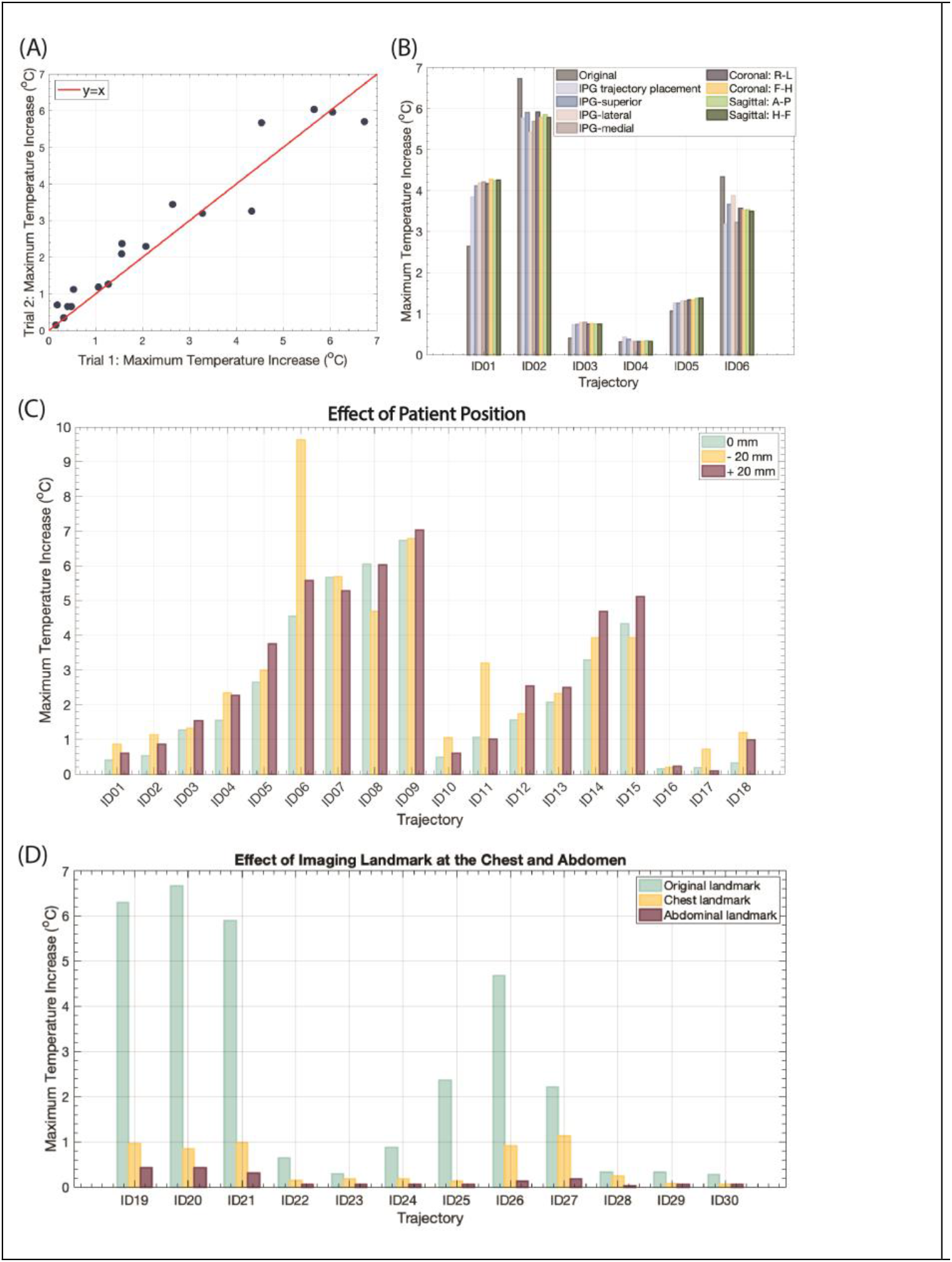
(A) ΔT_max_ from the test-retest experiments (ICC=0.96 with a 95% confidence interval of 0.91-0.99). (B) The effect of IPG configuration and imaging plane on ΔT_max_. (C) The effect of imaging position within the brain on ΔT_max_. The original imaging landmark corresponded 0 mm. (D) The effect of the imaging landmark at the chest and abdomen on ΔT_max_.

### Sensitivity analysis

Changing the patient position in the brain by ± 20 mm produced minimal differences in ΔT_max_. The mean ± standard deviation in |ΔT_max2_-ΔT_max1_| was 0.52 ± 0.98 °C, where ΔT_max1_ corresponded to the ΔT_max_ value when the phantom was at the central position, and ΔT_max2_ occurred when the phantom was superior or inferior to the central position. Figure 11. C. shows the ΔT_max_ at three brain imaging locations.

Altering the imaging landmark from the brain to the chest and the abdomen resulted in minimal RF heating around the DBS lead-tip. The mean ± standard deviation of ΔT_max_ was 0.49 ± 0.43 °C and 0.16 ± 0.15 °C when imaging was performed at the chest and abdominal landmarks, respectively (shown in Fig. 11. D). On the other hand, the mean ± standard deviation of ΔT_max_ was 2.58 ± 2.58 °C for the same lead trajectories when the experiments were conducted with the landmark located at the level of the DBS lead-tip. Thus, low-heating lead trajectories could mitigate RF heating regardless of the imaging landmark.

Similarly, altering the location of the IPG or the trajectory of the extension around the IPG had minimal effects on ΔT_max_ (shown in Fig. 11. B). The mean ± standard deviation in |ΔT_max2_-ΔT_max1_| was 0.66 ± 0.53 oC across all IPG-related configuration changes where ΔT_max1_ corresponded to the original configuration of the IPG. Furthermore, changing the slice acquisition and the phase encoding direction did not affect ΔT_max_, where the mean ± standard deviation of |ΔT_max2_-ΔT_max1_| was 0.66 ± 0.53 °C and ΔT_max1_ corresponded to axial slice acquisition with phase encoding in the anterior to posterior direction (shown in Fig. 11. B).

### Effectiveness of surgically implemented modified trajectories

Based on the findings from the phantom experiments, DBS lead trajectories with 2-3 concentric loops near the burr hole were implemented in six new patients (shown in Fig. 7. D). These patient-derived trajectories were replicated in phantom experiments with the Abbott DBS system, resulting in a mean ± standard deviation of ΔT_max_ of 1.24 ± 0.31 °C with a range of 0.87-1.57 °C. We compared the RF heating of DBS leads with surgically modified trajectories to six unmodified lead trajectories previously implemented in six other patients by the same neurosurgeons. The mean ± standard deviation of ΔT_max_ was 4.22 ± 0.57 °C with a range of 2.46-4.21 °C for the unmodified trajectories (shown in Fig. 12), demonstrating almost a 4-fold reduction in RF heating. Modifying the lead trajectory also reduced the variance of RF heating; a Levene’s test demonstrated that the variances between the modified and unmodified trajectories are unequal (p = 0.57).

**Figure 12:**
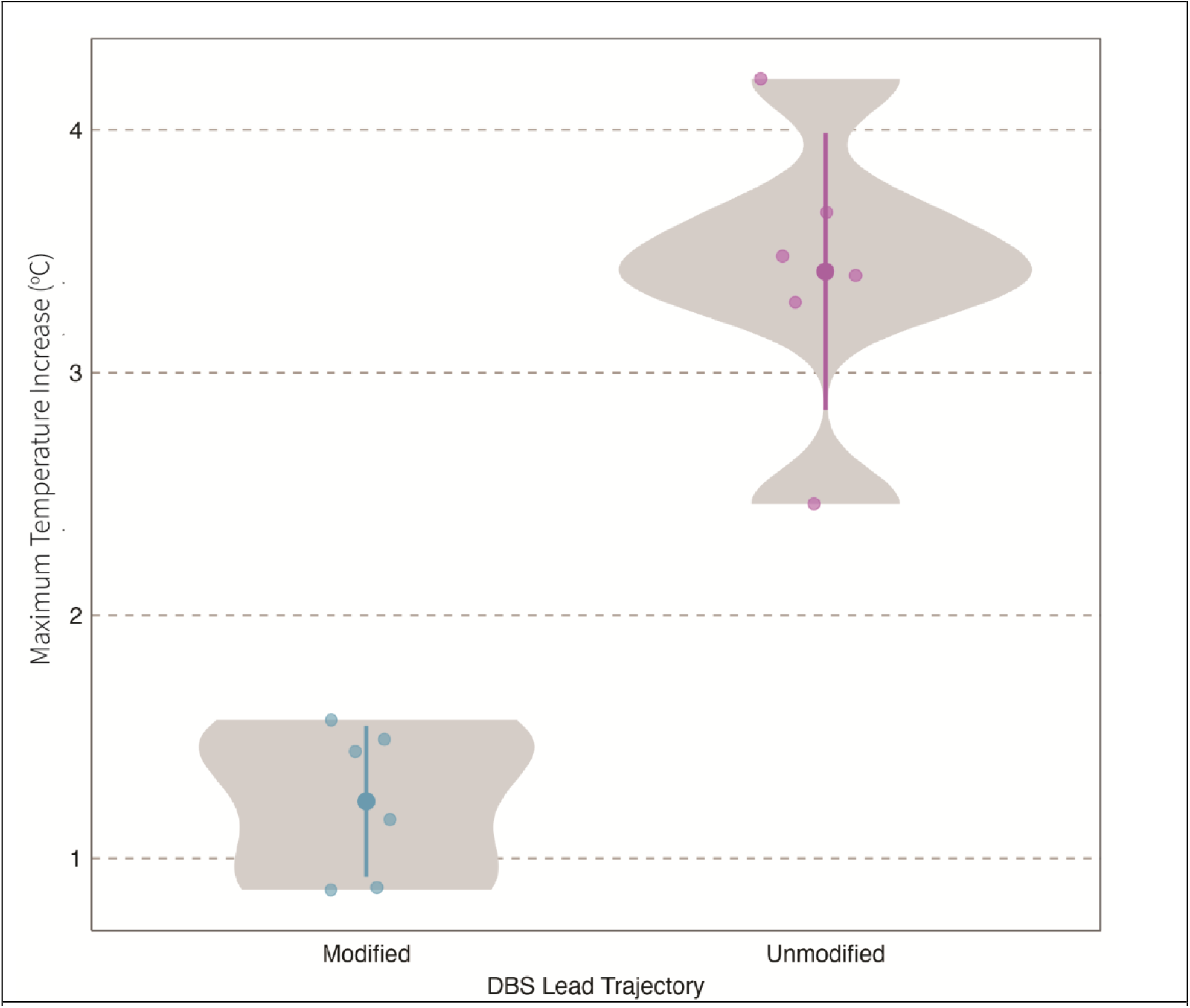
ΔT_max_ for the surgically modified and unmodified lead trajectories. There was almost a 4-fold reduction in the mean ΔT_max_ between unmodified and modified lead trajectories.

## Discussion/Conclusion

The past few years have witnessed a steady rise in the application of modern neuroimaging techniques for guiding and interpreting the outcomes of DBS therapy [48]–[51]. Although the majority of DBS devices are approved for MRI at 1.5 T under certain conditions, there is a scarcity of approved devices for 3 T MRI. However, there are strong incentives to work toward making 3 T MRI compatible with DBS. 3 T MRI confers a much better contrast-to-noise ratio, which makes it easier to delineate small abutting structures that can be crucial to localizing electrodes in areas like the subthalamic nucleus (STN) [52]. Furthermore, highly accelerated sequences with improved sensitivity that make use of functional MRI desirable for DBS are only accessible at 3 T [53]. Finally, while 3 T MRI is generally more sensitive to susceptibility effects, even this can act as an advantage when localizing electrodes in iron-rich areas of the brain (e.g., the STN) [54], [55] and for revealing pallidofugal and striatonigral fiber tracts [56].

The main barrier to MRI at 3 T for patients with implanted DBS systems is potential RF-induced heating around the lead-tip. It is well recognized that the trajectory of elongated conductive implants—such as leads encountered in neuromodulation and cardiovascular implantable electronic devices—has a significant effect on MRI-induced RF heating [57]–[60]. Safety studies on the RF heating of DBS systems have traditionally evaluated trajectories that elicit the worst-case scenario heating or looped trajectories without specificity [39]-[41]. Here, we present the first large-scale study to evaluate how characteristics of the extracranial DBS lead trajectory affect RF heating and to quantify the extent of RF heating reduction by surgical modification of the lead trajectory. Effective trajectory characteristics were readily implemented in new patients undergoing DBS surgery.

We found that placing 2-3 concentric, overlapping loops specifically within 40 mm of the surgical burr hole was most effective for reducing RF induced heating. Our results were consistent with the earliest introduction of a DBS lead management prototype that formed 2.25 successive loops ranging from 1.8 to 2.3 cm in diameter near the burr hole [38]. Other *in vitro* studies have incorporated loops in the DBS lead trajectories including a single 3 cm loop [43], 1-3 counterclockwise loops fixed on to the external surface of the phantom [39], 2 loops on the external surface of the phantom [40], and a single loop around the burr hole [41]. While all these trajectories can be classified as having looped topologies, their geometries and positioning were still vastly different. This present study provided a direct comparison to determine the characteristics of the trajectory that most effectively reduced RF heating; it is important to increase the number of concentric, overlapping loops near the surgical burr hole, but changing the size of the loops did not yield the same effect. These results also made it possible to refine recommendations on how to create low-heating trajectories that can be easily implemented.

Additional experiments were performed after perturbing the imaging landmark, IPG-related configurations, and the imaging plane to determine their effect on RF heating and if low-heating trajectories were resilient to such changes. The location of the imaging landmark is known to affect RF heating, and multiple patient positions in the head region are possible for clinical neuroimaging. However, changing the position in the head had a minimal effect on RF heating and changing the imaging landmark resulted in consistently lower ΔT_max_ across low, average, and high heating trajectories. Translating the position of the IPG in the superior, medial, and lateral directions also induced minimal changes in ΔT_max_ from the original configuration. Lastly, changing the slice acquisition and phase encoding direction had a minimal effect on ΔT_max_. This is expected as the specific slice and the encoding directions correspond with the MRI gradient coils which do not produce gradient-induced heating for medical devices with elongated leads such as a DBS system.

Our preliminary clinical results demonstrated that modifying the extracranial DBS lead trajectory is feasible within the current surgical procedure without increasing the complexity or duration. The modified DBS lead trajectories reduced RF heating during 3 T MRI by almost 4-folds compared to the unmodified lead trajectories previously implanted by the same neurosurgeons. The variances of ΔT_max_ between the implanted leads with modified versus unmodified trajectories are unequal and implementing the modified trajectories can make the RF heating more predictable. This demonstrates great potential for widespread adoption of surgical modification of the lead trajectory as both neurosurgeons in this study presented different surgical practices to produce the concentric looped trajectories.

Surgically modifying the extracranial DBS lead trajectory while focusing on increasing the number of concentric loops and the loops’ placement can effectively mitigate RF heating during 3 T MRI. The reduction in RF heating was most apparent for leads that were contralateral with respect to the IPG as the parameter space is larger. Clinical adoption of the trajectory specifications was feasible, and subsequent experiments confirmed low RF heating. Overall, this method can enable safer imaging during MRI at 3 T for patients with DBS systems.

## Data Availability

All data produced in the present study are available upon reasonable request to the authors

