## Supplementary Information for "Surgical modification of deep brain stimulation lead trajectories substantially reduces RF heating during MRI at 3 T: From phantom experiments to clinical applications"

**Supplementary Material**

To construct an anatomically representative skull, we first compared the size of the skull to the head dimensions of patients with DBS systems. We measured the maximum head breadth, maximum head length, and the distance from the most superior point on the head to an axial plane intersecting the eyebrows from patient computed tomography (CT) images (shown in Fig. S1). The maximum head breadth, length, and distance of the skull was 141 mm, 185 mm, and 91 mm, respectively. Figure S2 shows the distribution of the maximum head breath, maximum head length, and the distance between the eyebrows and the most superior point on the head across 53 patients. The mean ± standard for the maximum breadth, length, and distance was 157.3 ± 8.7 mm, 198.9 ± 11.6 mm, and 100.0 ± 11.4 mm, respectively.

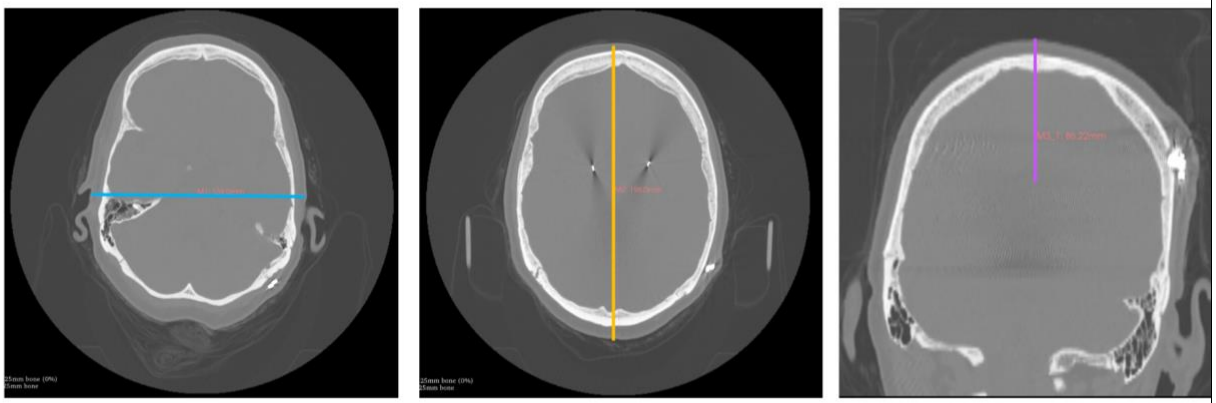

**Figure S1:** Example measurements of the maximum head breadth (left), maximum head length (center) and the maximum distance between the most superior point on the head and the level of the eyebrows (right) as indicated by the blue, orange, and purple lines, respectively. All measurements were performed by evaluating patient CT images in 3D Slicer.

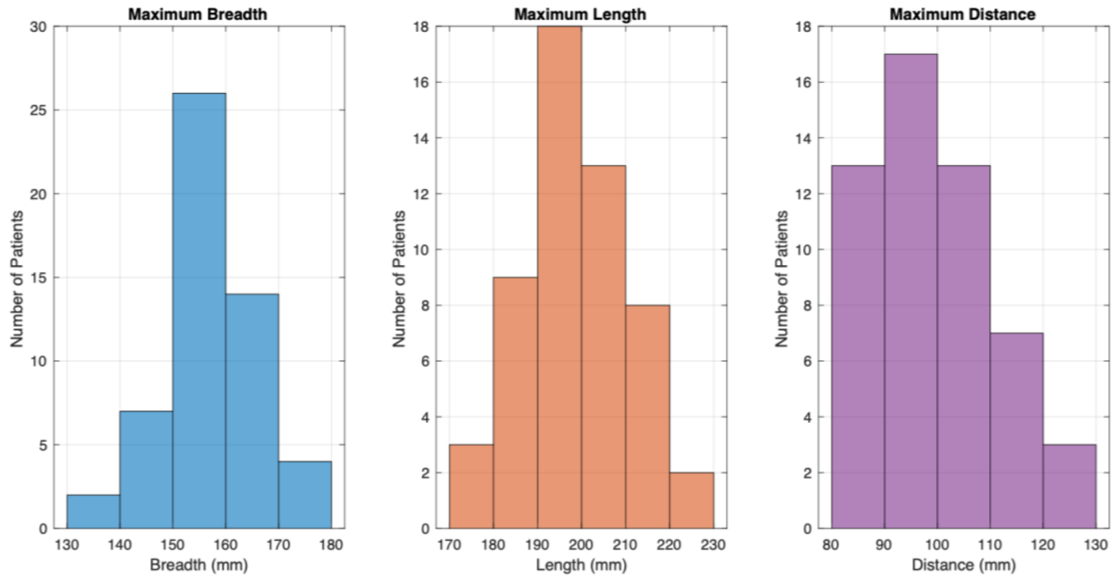


**Figure S2:** The distribution of the maximum head breath (left), maximum head length (center), and the maximum distance between the eyebrows and the most superior point on the head (right) across 53 patients.
